# Perceived seizure risk in epilepsy – Chronic electronic surveys with and without concurrent EEG

**DOI:** 10.1101/2023.03.23.23287561

**Authors:** Jie Cui, Irena Balzekas, Ewan Nurse, Pedro Viana, Nicholas Gregg, Philippa Karoly, Gregory Worrell, Mark P Richardson, Dean R. Freestone, Benjamin H. Brinkmann

## Abstract

**Objective:** Previous studies suggested that patients with epilepsy might be able to forecast their own seizures. We sought to assess the relationships of premonitory symptoms and perceived seizure risk with future and recent self-reported and EEG-confirmed seizures in the subjects living with epilepsy in their natural home environments.

**Methods:** We collected long-term e-surveys from ambulatory patients with and without concurrent EEG recordings. Information obtained from the e-surveys included medication compliance, sleep quality, mood, stress, perceived seizure risk and seizure occurrences preceding the survey. EEG seizures were identified. Univariate and multivariate generalized linear mixed-effect regression models were used to estimate odds ratios (ORs) for the assessment of the relationships. Results were compared with device seizure forecasting literature using a mathematical formula converting OR to equivalent area under the curve (AUC).

**Results:** Sixty-nine subjects returned 12,590 e-survey entries, with four subjects acquiring concurrent EEG recordings. Univariate analysis revealed increased stress (OR = 2.52, 95% CI = [1.52, 4.14], *p <* 0.001) and decreased mood (0.32, [0.13, 0.82], 0.02) were associated with increased relative odds of future self-reported seizures. On multivariate analysis, previous self-reported seizures (4.24, [2.69, 6.68], *<* 0.001) were most strongly associated with future self-reported seizures, and high perceived seizure risk (3.30, [1.97, 5.52], *<* 0.001) remained significant when prior self-reported seizures were added to the model. No significant association was found between e-survey responses and subsequent EEG seizures.

**Significance:** It appears that patients may tend to self-forecast seizures that occur in sequential groupings. Our results suggest that low mood and increased stress may be the result of previous seizures rather than independent premonitory symptoms. Patients in the small cohort with concurrent EEG showed no ability to self-predict EEG seizures. The conversion from OR to AUC values facilitates direct comparison of performance between survey and device studies involving survey premonition and forecasting.

**Key points:**

- Long-term e-surveys data and concurrent EEG signals were collected across three study sites to assess the ability of the patients to self-forecast their seizures.
- Patients may tend to self-forecast self-reported seizures that occur in sequential groupings.
- Factors, such as mood and stress, may not be independent premonitory symptoms but may be the consequence of recent seizures.
- No ability to self-forecast EEG confirmed seizures was observed in a small cohort with concurrent EEG validation.
- A mathematic relation between OR and AUC provides a means to compare forecasting performance between survey and device studies.

## 1 Introduction

Although seizures generally occupy a relatively small portion of absolute time of a large majority of people with epilepsy, the unpredictability of seizures is an overwhelming challenge of living with epilepsy [1]–[4], the ability to forecast seizures may have a significant impact on patients’ everyday lives[4], [5]. Seizure forecasting has been established using intracranial or scalp electroencephalography (iEEG/EEG)[5], [6]. More recently, research on forecasting have sought to use wearable devices[7] to identify factors or premonitory symptoms associated with seizures in order to provide patients with noninvasive tools to better manage their epilepsy[8]–[10]. Previous studies have demonstrated that some patients experienced premonitory symptoms, or a sense of elevated seizure risk, before seizures[9]–[13], though which patients were strong predictors and how reliably they predicted their seizures varies. Seizure self-forecasting studies have primarily relied on electronic, self-reported seizure diaries to record seizure events, although these records can be unreliable for many patients when validated with EEG[6], [14], [15].

An important methodological consideration in seizure forecasting is whether predictive factors are validated with EEG-confirmed seizure. Without concurrent EEG recordings, it is difficult to determine if a behavioral event is a true seizure associated with an ictal electrographic. Additionally, many EEG seizures are missed if patients are amnestic to or unaware of the events[6], [16], [17]. Ultra-long term (months to years) studies with concurrent iEEG/EEG monitoring were technically unavailable until recently, and has been limited mostly to experimental implanted devices[18]–[20]. However, reliable technologies for long-term monitoring (both invasive[6], [20] and non-invasive[21]) are emerging and are being used to explore brain activity[20], [22], [23] and peripheral physiology to aid seizure forecasting[7], [24].

Furthermore, it is difficult to assess the relative forecasting performance of implanted or wearable devices to the reported results of diary-based self-forecasts, as different performance metrics are used in the respective literature. Area under the curve (AUC) of the receiver operating characteristic (ROC) curve and time in warning (TIW)[25] are commonly used in device studies, while odds ratios (OR) computed from statistical models[10], [26], [27] are often used in the seizure diary self-forecasting literature. The ability to directly compare these different metrics would be particularly useful[28] in assessing whether a device may offer a patient meaningfully better seizure forecasts than their own premonitory intuition.

In this study we investigated long-term electronic survey (e-survey) data from people living with epilepsy, some of whom had concurrent long-term EEG monitoring devices. Acquisition of survey data occurred across three international sites and in conjunction with wearable and implanted device recordings where possible. We assessed the relationships of premonitory symptoms and perceived seizure risk with future and recent self-reported and EEG-confirmed seizures in a cohort of people living with epilepsy in their natural home environments. In addition, we sought to compare the accuracy of seizure self-forecasts using perceived seizure risk and/or premonitory symptoms to published data using noninvasive and invasive devices.

## 2 Methods

### 2.1 Subject recruitment

This is a multi-center study with data collected from Seer Medical (Melbourne, Victoria, Australia), Mayo Clinic (Rochester, MN, USA), and King’s College London (London, UK). Subjects were recruited across the three sites from among patients with epilepsy as part of the My Seizure Gauge project (Clinical Trials: NCT03745118)[7], [21], [29]. Subjects participating in the electronic surveys with a smartphone application (Seer Medical™)[30] without concurrent EEG devices were recruited by Seer Medical (Seer cohort). In addition, subjects participating in the surveys from Mayo Clinic with concurrent intracranial EEG (iEEG) recording (NeuroPace RNS^®^) were recruited (Mayo cohort). Additional subjects were recruited at KCL (KCL cohort), who had a CE-marked implantable subcutaneous EEG (sqEEG) system[20] (UNEEG SubQ™, Lillerod, Denmark). The e-survey data provide a direct comparison to previously published work[10], [11], [13], while the concurrent EEG signals provide an opportunity to assess the relationship between self-reported seizures and EEG-confirmed seizures.

All activities were approved by St. Vincent’s Hospital Melbourne Human Research Ethics Committee at Seer Medical, Mayo Clinic IRB: 18-005483 “Human Safety and Feasibility Study of Neurophysiologically Based Brain State Tracking and Modulation in Focal Epilepsy”, and Institutional Review Boards of Ethics Committees at King’s College London, and all subjects provided informed consent.

### 2.2 Data acquisition

#### 2.2.1 E-survey data

All participants were instructed to conduct surveys using Seer mobile application[31] (Seer Medical, Melbourne, Australia[30]). No restriction on the time and number of surveys they could complete was imposed, although they were asked to participate daily (Figure 1). Except one subject at KCL, all other subjects completed the same set of six (6) survey questions, as follows (see Supplemental Table 1):

**Figure 1:**
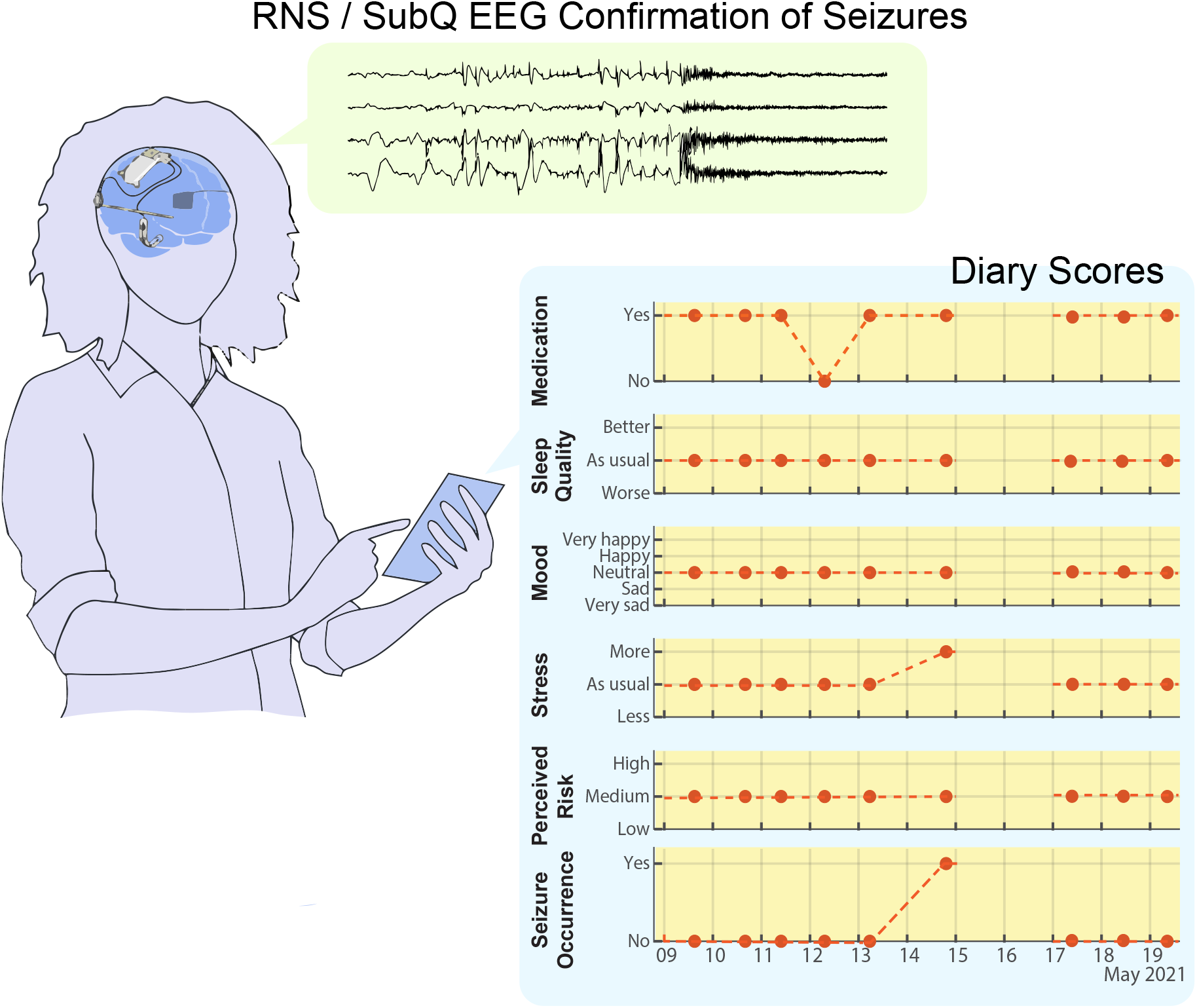
Schematic diagram of data acquisition: Electronic surveys were conducted with a mobile smartphone application and consisted of six questions to assess medication compliance, sleep quality, mood, stress, perceived risk, and seizure occurrence. A subset of subjects had the cranially implanted Neurospace™ RNS EEG device or the UNEEG™ subcutaneous EEG (sqEEG) device for concurrent EEG recordings to provide EEG-validated seizure diaries. Abbreviation: RNS, responsive neurostimulator; KCL, King’s College Long; SubQ EEG, subcutaneous encephalography.

1. *Medication compliance*. Medication compliance was assessed by the survey question: “Did you take all of your prescribed epilepsy medication in the last 24 hours?” with the response choices of “Yes”, “No”, or “Not Available”.
2. *Sleep quality*. Subjects assessed the sleep quality before the survey (“How did you sleep last night compared to usual?”) on a three-point scale: “Worse”, “As usual”, or “Better”.
3. *<u>Mood</u>*. Subjects rated their level of mood (“Rate your current mood.”) on a five-point scale: “Very sad”, “Sad”, “Neural”, “Happy”, and “Very happy”.
4. *<u>Stress</u>*. Subjects rated their level of stress (“Are you more or less stressed than you usually are?”) with three responses: “Less”, “As usual” and “More”.
5. *<u>Perceived risk</u>*. The patient’s perceived risk of future seizures was assessed by the question: “What do you feel your risk of a seizure is in the next 24 hours?” with a choice of three responses: “Low”, “Medium”, and “High”.
6. *<u>Seizure occurrence</u>*. Finally, the survey asked if the subject had a seizure before completing the survey (“Have you had a seizure in the last 24 hours?”) with the choice of either “Yes” or “No”. One subject at KCL used a slightly different set of questions in the survey due to a software upgrade to the mobile application during the study. We converted the responses of this subject to match the responses of the other set of questions (see Supplemental Table 1). Note that since the seizure occurrence question was not applied to this subject, we retrieved this information from the self-reported diaries recorded with Seer mobile application.

#### 2.2.2 EEG-confirmed seizures

We acquired two types of EEG signals from the subjects with concurrent EEG. Subjects at KCL were implanted with the UNEEG™ SubQ devices. The recording of the continuous sqEEG signals has been described previously[20], [22], [32]. Briefly, the minim-invasive sqEEG device was implanted unilaterally under local anesthesia, over the region of pre-identified ictal EEG changes, recording 2-channel bipolar EEG at a sampling rate of 207 Hz. Offline, the seizure events were identified with UNEEG commercially available software and reviewed by an epileptologist with experience in subcutaneous EEG (P.V.).

Similarly, iEEG signals were acquired from the two subjects at Mayo equipped with NeuroPace™ RNS devices[18], [33], [34]. The RNS device stored up to 12 minutes of iEEG signals from the detected epileptiform activity. Each candidate EEG seizure in the recording was assessed by two expert reviewers, one of whom is a board-certified epileptologist (G. W.). A lead EEG seizure was identified as a seizure which had onset at least 4 hours away from the onset of the preceding seizure.

#### 2.2.3 Seizure events

Variables were analyzed with respect to self-reported and EEG-confirmed seizures, with attention given to the timing of events relative to the survey responses.

- *Reported seizure in prior / next 24 hours*: For each time instance of a survey response, we reviewed reported seizure occurrences to determine whether a seizure was reported in the 24 hours before the survey, and whether a seizure was reported in the 24 hours following the survey response (future reported seizure).
- *EEG seizure in prior / next 24 hours*: At each instance of the electronic survey, we determined whether an EEG seizure occurred within 24 hours before the survey, and whether an EEG seizure occurred within 24 hours after the survey (future EEG seizure).

### 2.3 Statistical analysis

#### 2.3.1 Odds Ratio (OR)

Univariate and multivariate generalized linear mixed effect regression (GLMER) models (logit-normal with random subject specific intercepts to take into account within subject correlation) were used to calculate ORs[26] as previously described[10]. For e-survey data of all subjects, Jackknife[35] standard errors were used to estimate approximate p-values and 95% confidence intervals (CIs). Due to the limited number of subjects with concurrent EEG the Jackknife approach was inappropriate. Instead we resorted to the covariance matrix of the GLMER model (see R function *glmer* in package lme4[36]) for the estimation of standard errors and then approximate *p*-values and 95% CIs. We examined the predictors (independent variables) as continuous variables. The estimated OR reflected the relative change of odds of the responses (dependent variables) as a function of a unit change of the predictors.

#### 2.3.2 Positive predictive value (PPV)

A PPV[37] was adopted to estimate approximately the probability (or frequency) of reported seizures in addition to the ORs.

#### 2.3.3 Relationship Between Odds Ratio (OR) and Area Under the Curve (AUC)

The OR has been widely accepted as a single indicator of diagnostic performance[26], while AUC has been preferred in device studies to evaluate the performance of seizure detectors. To directly compare performance from the present study to those previously reported device studies, we propose a technique to convert OR to AUC. Specifically, we derived an equal OR curve (Equation 1) on the FPR (false positive rate) - TPR (true positive rate) plot, under the condition of constant OR for all possible cutoff values,

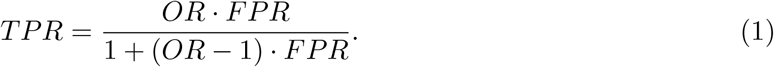

The relationship between OR and AUC can then be found (see Appendix A for the derivation) as,

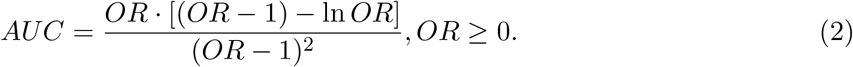

Note that Glas et al.[26] proposed a similar relationship, but without a closed form equation.

## 3 Results

Sixty-nine subjects returned 12,590 total e-survey entries (see Supplemental Table 2) with 157 ± 23 (mean ± SD) entries per subject across the three sites. Reported seizure rate is shown in Figure 2A. A summary of demographic data is presented in Table 1.

**Figure 2:**
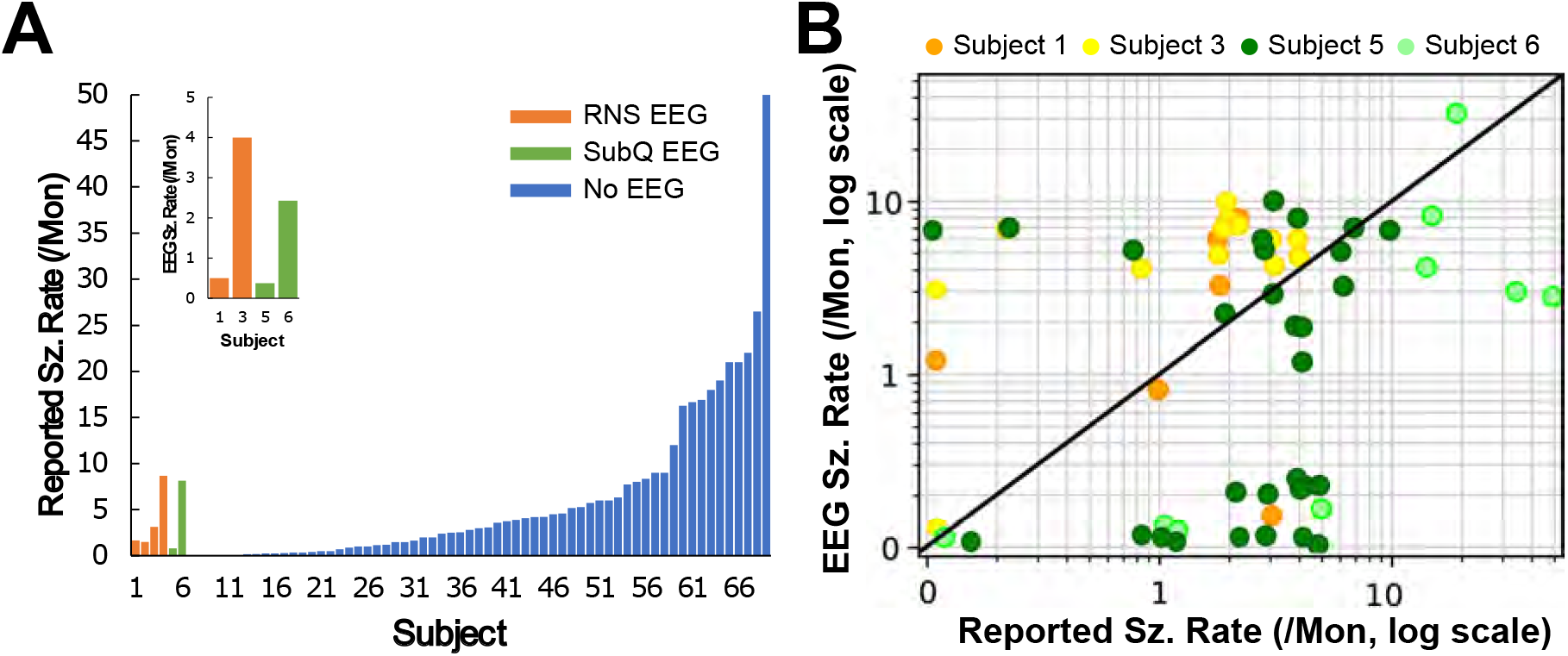
Reported seizure and EEG seizure rates. (A) Reported and EEG seizure rates (events/month). The main graph shows reported seizure rates of all 69 subjects (SOG, n = 69), where the inset shows the EEG seizure rate of the four subjects with reliable EEG seizure detections (CEG, n = 4). Blue bars indicate subjects without concurrent EEG (n = 63), green with subscalp EEG (n = 2), and orange with the Neuropace RNS (n = 4 for reported seizures and n = 2 for EEG seizures); (B) Scatter plot of reported and EEG seizure rates (SOG, n = 4). Each dot indicates the estimated rates of reported and EEG seizures in one month. Orange-shade dots are subjects with the Neuropace™ RNS device (Subject 1 & 2) and green-shade with the UNEEG™ SubQ device (Subject 5 & 6). The dashed diagonal line indicates identity between reported and EEG seizure rates. Abbreviation: Sz., seizures; SubQ EEG, subcutaneous encephalography; Mon: Month.

**Table 1:** Characteristics of demographic data. † Partial subjects from Seer cohort chose not to disclose either gender or age, or both. †† Information about epilepsy types of some subjects from Seer cohort is not available. ††† Only from subjects with concurrent EEG recordings.

| **Characteristic** |  |
| --- | --- |
| **Gender** | *No. of subjects (%)* |
| Male | 17 (24.6) |
| Female | 33 (47.8) |
| Not available† | 19 (27.5) |
| **Age [year]†** | *Median* |
| Male | 58 |
| Female | 37 |
| All | 44 |
| **Epilepsy Type** | *No. of subjects (%)* |
| Focal | 27 (39.1) |
| Generalized | 12 (17.4) |
| Not available†† | 30 (43.4) |
| **Concurrent EEG†††** | *No. of subjects* |
| Temporal | 3 |
| Hippocampus | 3 |

Of the 69 subjects who returned at least 50 e-survey responses, 50 disclosed either gender or age, or both. The range of reported ages was 18 - 78 years. Thirty-nine subjects had information on seizure localization. Additional information about individual subjects is included in Supplemental Table 3^1^. Four subjects in Mayo cohort with the RNS device returned an adequate number of survey responses for inclusion, and of these it was found that two subjects’ devices were triggering more long-episode candidate seizure detections than could be stored on the device, rending their EEG seizure records severely limited. These subjects were included in the e-survey and spike rate analyses but were excluded from the EEG seizure analyses. The two subjects from KCL cohort returned adequate e-survey responses and were included in all analyses.

The 69 subjects providing e-survey data were defined as survey only group (SOG, n = 69), while the four subjects providing additional EEG recordings as concurrent EEG group (CEG, n = 4).

### 3.1 Perceived risk significantly associated with future reported seizure

#### 3.1.1 Univariate analysis

We analyzed ORs between premonitory factors (stress, mood, sleep quality and medication) and reported seizure in next 24 hours using a univariate regression analysis of SOG (n = 69) (Figure 3A). Stress and mood were significantly associated with future reported seizure. A one unit increase in stress was associated with relative odds of 2.52 of reported seizure in next 24 hours (95% CI = [1.53, 4.14], p *<* 0.001), indicating higher stress correlated with higher probability of following reported seizure. Moreover, a unit change in mood was associated with relative odds of 0.32 (95% CI = [0.13, 0.82], p = 0.02), indicating lower mood correlated with increased probability of reporting a seizure in next 24 hours. No significant associations were observed with sleep quality (p = 0.30) and medication (p = 0.36).

**Figure 3:**
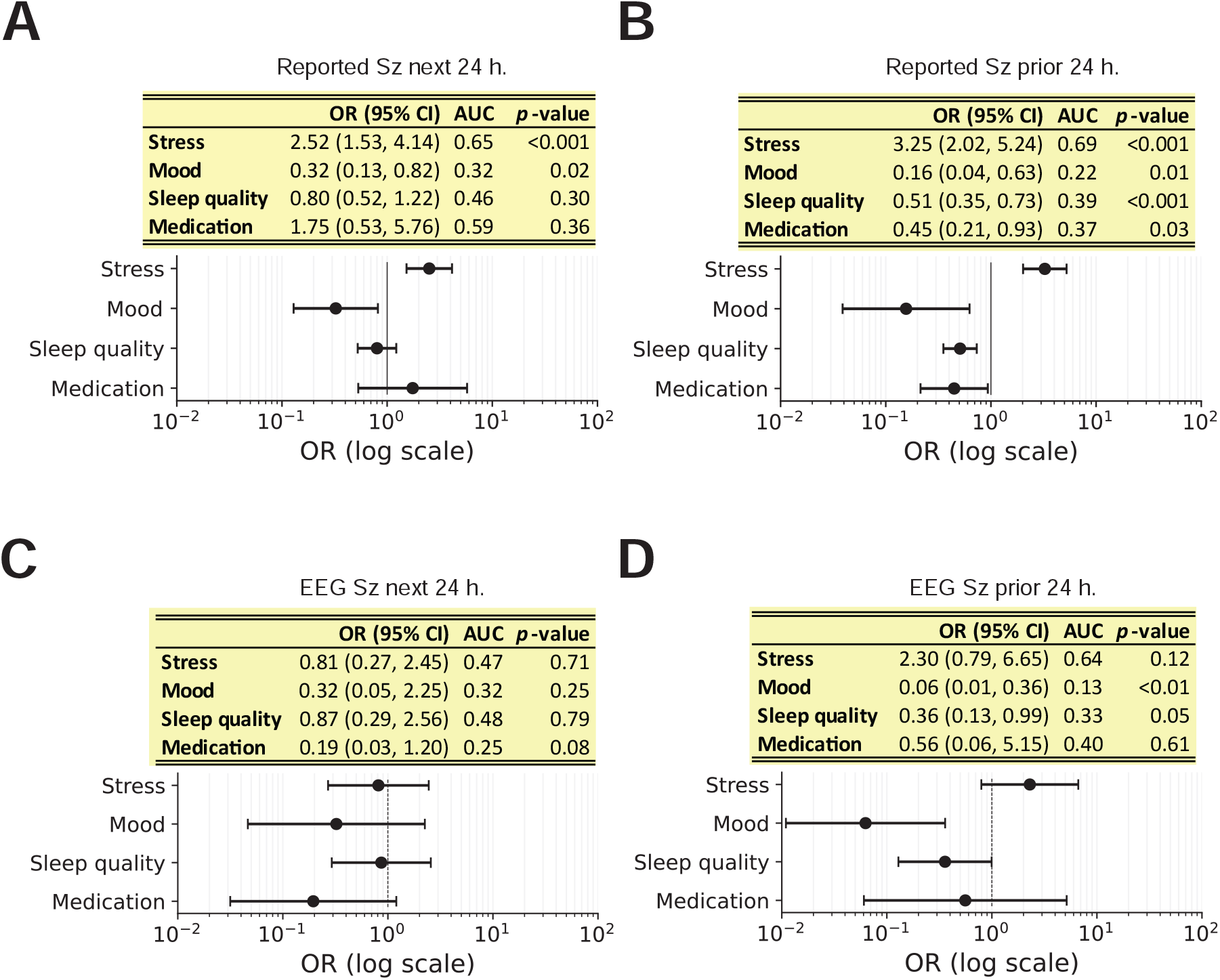
Univariate regression analysis in survey only group (SOG, n = 69) (A, B) and concurrent EEG group (CEG, n = 4) (C, D). In each panel, the title describes the response variable. The table presents the estimated values of OR, 95% CI, AUC and p-value corresponding to the error bars shown below. ORs between the premonitory factors (medication, sleep quality, mood and stress) and (A) reported seizures in next 24 hours, (B) reported seizures in prior 24 hours, (C) EEG seizure in next 24 hours and (D) EEG seizure in prior 24 hours are shown. Note that the standard errors of ORs in SOG were estimated with Jackknife method, while standard errors in CEG were estimated from covariance matrixes of the models (see text for the details). Abbreviation: OR, odds ratio; Sz, seizure; h., hour; CI, confidence interval; AUC, area under the curve.

To assess the potential influence of seizure grouping or clustering on seizure reports, we analyzed ORs between survey responses and reported seizure in prior 24 hours (Figure 3B). We found that all four factors showed significant associations. Increased stress (OR = 3.25, 95% CI = [2.02, 5.24], p *<* 0.001), decreased mood (0.16, [0.04, 0.63], 0.01), lower sleep quality (0.51, [0.35, 0.73], *<* 0.001) and lower medication compliance (0.45, [0.21, 0.93], 0.03) were each significantly correlated with an increased probability of having reported a seizure in the prior 24 hours before the survey.

#### 3.1.2 Multivariate analysis

To assess for possible confounds and interactions among premonitory factors, we performed a multivariate regression analysis to estimate the ORs between the survey responses and reported seizures in the next 24 hours (Figure 4). In Model 1 (Figure 4A) stress, mood, sleep quality and medication were included as predictors. As in the univariate analysis (Figure 3A), a unit increase in stress was significantly associated with relative odds of 2.15 of reported seizure in next 24 hours (95% CI = [1.25, 3.70], p = 0.01). Unlike the univariate analysis, however, mood (p = 0.24) was no longer significantly associated with future seizure report.

**Figure 4:**
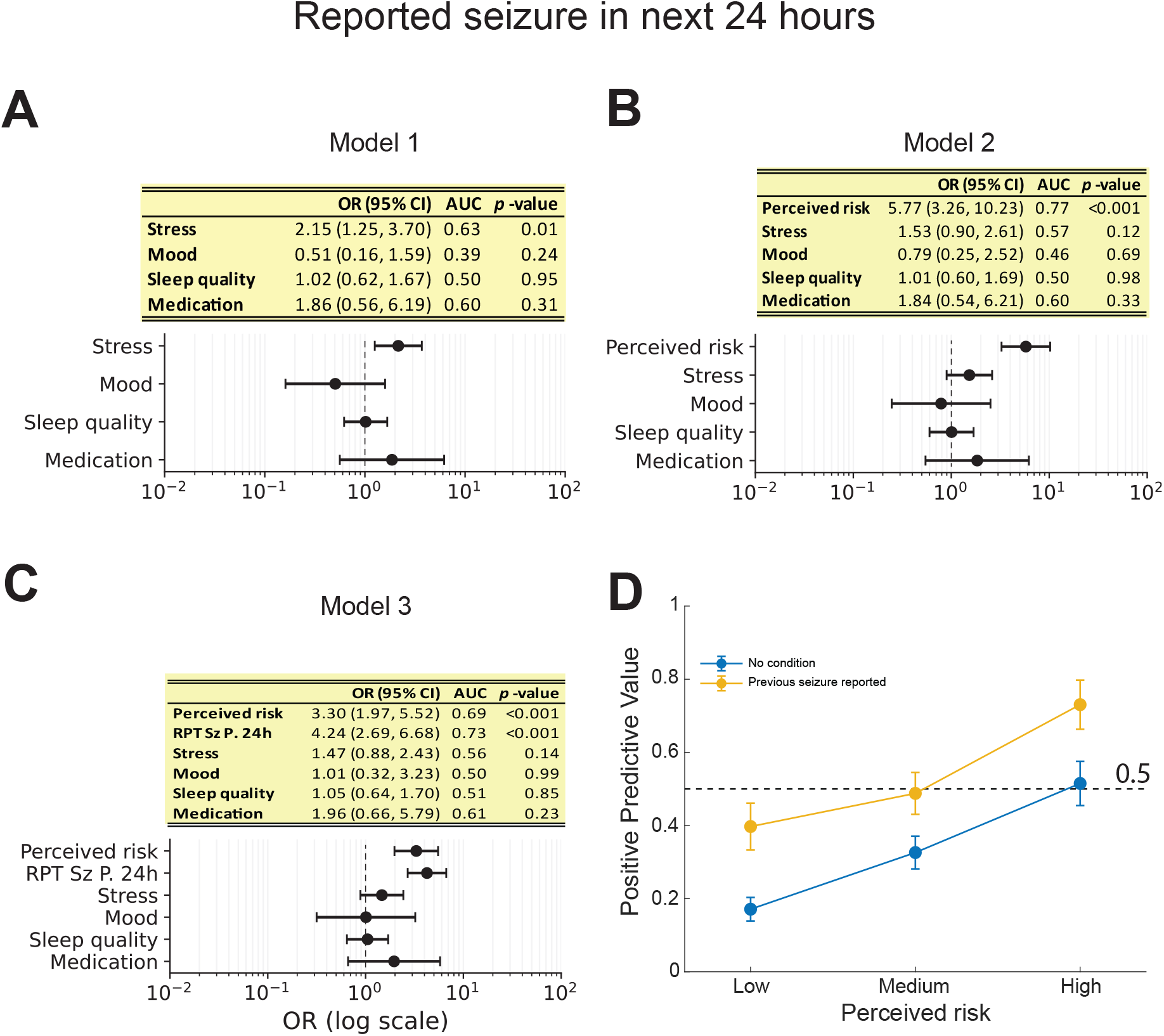
Multivariate regression modeling and positive predictive value (PPV) analysis of reported seizure in next 24 hours in survey only group (SOG, n = 69). OR analyses are shown in (A) - (C). In each panel, the table presents the estimated values of OR, 95% CI, AUC and p-value (Jackknife standard error), corresponding to the error bars shown below. ORs corresponding to the combination of a set of predictors of (A) premonitory factors (medication, sleep quality, mood and stress) only (Model 1), or (B) premonitory factors plus perceived risk (Model 2), or (C) reported seizure in prior 24 hours besides premonitory factors plus Perceived risk (Model 3), are shown; (E) PPV of reported seizure in next 24 hours at three levels of perceived risk (Low, Medium and High). Blue dots indicate the PPVs independent on previous seizure report, while yellow dots the PPVs given that seizures were reported in prior 24 hours. PPV Error bar: ±1 SEM. Abbreviation: OR, odds ratio; CI, confidence interval; RPT, reported; Sz., seizure; P., prior; h.; hours; AUC, area under the curve; SEM, standard error of mean.

When perceived risk was included in regression Model 2 (Figure 4B), neither stress (p = 0.12) nor mood (p = 0.69) remained significantly associated with future reported seizures. Only perceived risk of seizure in the next 24 hours was significantly associated with reported seizures in the next 24 hours (OR = 5.77, 95% CI = [3.26, 10.23], p *<* 0.001), which implies interdependence among perceived risk, stress and mood.

Many patients experience clusters or groups of seizures[38], [39]. To evaluate the potential influence of seizure clustering on seizure reporting, we further included reported seizures in the prior 24 hours in Model 3 (Figure 4C). We found that both perceived risk and reported seizures in the prior 24 hours were significantly associated with reported seizures in next 24 hours. Increased perceived risk was significantly associated with an increased probability of reporting a seizure in the following 24 hours (OR = 3.30, 95% CI = [1.97, 5.52], p *<* 0.001). Similarly, recently reported seizures showed a significant association with future reported seizures (4.24, [2.69, 6.68], *<* 0.001) on a 24-hour timescale. No other survey responses showed a significant association in Model 3. These results suggest that both perceived risk and recently reported seizures are both associated with future seizure reporting, with recently reported seizures showing a stronger association.

### 3.2 Association with EEG-confirmed seizures

#### 3.2.1 Weak correlation between reported and EEG seizures

We investigated the correlation between survey responses and EEG confirmed seizures in CEG (n = 4). EEG seizure rate of these four subjects is shown in the inset of Figure 2A. Total monthly reported and EEG seizures were weakly correlated (Pearson-correlation r between 0.17 and 0.33 for all participants), but the correlation was not statistically significant (all p ≥ 0.28, Figure 2B).

#### 3.2.2 Univariate analysis

We further analyzed the ORs between premonitory factors and EEG seizures in the univariate regression analysis of the CEG (n = 4) (Figure 3C, D). No significant associations were found with EEG seizures in next 24 hours (Figure 3C). We then evaluated the association with EEG seizures in the prior 24 hours (Figure 3D). Lower mood (0.06, [0.03, 0.15], *<* 0.001) and lower sleep quality (0.36, [0.13, 0.99], 0.05) were significantly associated with an increased probability of EEG seizures before the survey.

#### 3.2.3 Multivariate analysis

We also conducted multivariate regression modeling in the CEG to assess possible interdependence between factors (Figure 5). In this analysis, the predictors (independent variables) were perceived risk, EEG seizure in prior 24 hours, reported seizure prior 24 hours, stress, mood, sleep quality and medication. Using reported seizures in the next 24 hours as the dependent variable (Figure 5A), significant associations were found with both reported seizures in the prior 24 hours (OR = 6.46, 95% CI = [2.76, 15.11], p *<* 0.001), and stress (0.13, [0.02, 0.89], 0.04), with decreased stress associated with future seizure reports. No other predictor was found to have a significant association with future reported seizures. Medication compliance could not be reliably modeled in this analysis.

**Figure 5:**
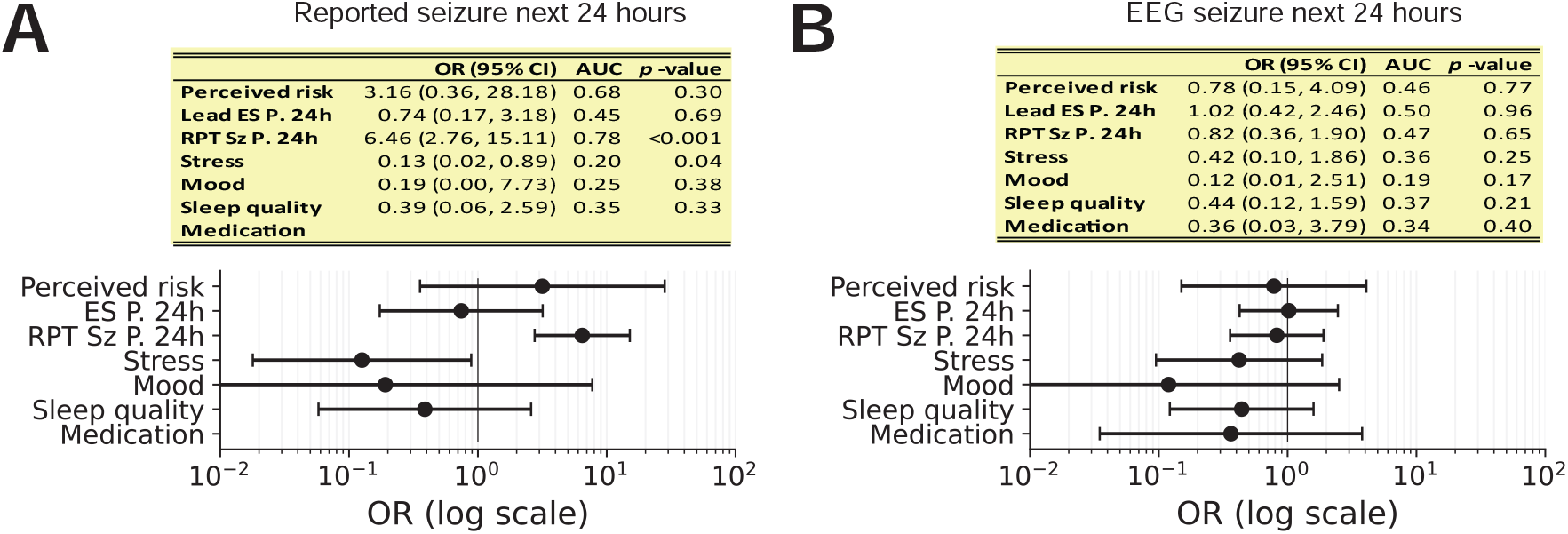
Multivariate regression modeling in concurrent EEG group (CEG, n = 4). ORs between the premonitory factors, reported seizures in the prior 24 hours, EEG seizures in the prior 24 hours and perceived risk in next 24 hours, and (A) self-reported seizures in next 24 hours, or (B) EEG seizures in next 24 hours. Note: standard errors were estimated from covariance matrix of the multivariate GLMER model; OR involving medication compliance could not be reliably estimated in (A). Error bar: 95% CI. Abbreviation: OR, odds ratio; CI, confidence interval; ES, EEG seizure; RPT, reported; Sz, seizure; P., prior.

For EEG seizures occurring in the next 24 hours, no predictors were found to be significantly associated (Figure 5B). Additional analyses investigated the ORs between average interictal spike rates (see Supplemental Materials A) measured prior to surveys using 3 hour, 24 hour, and the prior overnight periods and perceived seizure risk, reported seizures in the next 24 hours, reported seizures in the prior 24 hours, mood, medication, sleep quality and stress (Supplemental Figure 2). No significant associations were found with any factors.

### 3.3 Reporting seizures before the survey elevated the probability of future reported seizure

To further investigate the influence of recently reported seizures prior to the survey on future reported seizures, we analyzed ORs and PPV for reported seizure in next 24 hours in the SOG (n = 69), only for seizures with a previous seizure reported within 24 hours before the survey (Figure 4D, Supplemental Figure 1). We analyzed the PPV of perceived seizure risk for reported seizures following the survey (Figure 4D). Both PPV curves showed positive slopes as a function of scales of perceived risk, which were consistent with the estimated ORs being greater than 1. More importantly, a significant OR between perceived risk and reported seizure in next 24 hours did not translate to a high probability of future reported seizures. As shown in PPV without condition (blue error bars), even if the perceived risk was High, the PPV was only around 0.5, essentially a random guess. On the contrary, given previous seizure reported (yellow error bars), the PPV was elevated to ∼0.7.

### 3.4 Closed form equation between OR and AUC

Estimated AUC values from OR values throughout our tables and figures are reported to facilitate comparison to device performance (Supplemental Figure 3 illustrates a curve relating OR to the corresponding estimated AUC, as proposed in Equation (2), using an example OR of 3.5, which translates to an AUC of 0.70).

## 4 Discussion

Our results using electronic seizure surveys agree with prior literature that shows a significant association between perceived seizure risk and future self-reported seizures in the next 24 hours[9], [10], [12]. However, our results also show that the associations of mood and stress with future self-reported seizures do not persist when recent self-reported seizures are factored into the regression model, although perceived seizure risk does retain a significant association. This may suggest that low mood and increased stress are caused by recent seizures, and that the observed premonitory effects of these factors could be an artifact of seizures occurring in groups or clusters. Given this tendency for seizure clustering, one may wonder whether patients’ sense of elevated seizure risk could arise from simply knowing that their seizures tend to occur in groups. However the persistence of the significant association of perceived risk and future self-reported seizures when recent seizures are added to the regression model suggests this is not the case. The origin and nature of this effect remains to be explored further and validated with objective EEG confirmation of seizures in a large cohort. The OR of future reported seizures given elevated perceived seizure risk of 3.30 in the full multivariate regression (Model 3, Figure 4C) is similar to the findings in previous studies[12] and corresponds to an AUC of 0.69 using Equation 2 above. It may be reasonable to use the accuracy of self-forecasts as a lower bound to what would be considered acceptable in a forecasting device system, and recently published smartphone and wearable device studies have exceeded this benchmark[7], [21], [32], [40]. While this comparison may be reasonable when concurrent EEG validation is not possible, EEG-confirmed seizures should be the gold standard of device and survey studies alike. In concurrent EEG group, no ability to self-forecast seizures was apparent. Although the small cohort limited the statistical power of the comparison, it is notable that perceived risk’s association with self-reported seizures was nearly identical to the larger group’s association (OR 3.16 vs. 3.30), while the association with EEG-confirmed seizures in this group was 0.78.

The limited size of the concurrent EEG cohort makes it difficult to draw clear conclusions based on objectively measured seizures. Our small cohort showed the same difficulties in accurate self-reporting described previously[6], [16], [17], although there was more over-reporting of seizures in our cohort than has been apparent in previous studies (Figure 2). In the univariate analysis low mood was associated with both self-reported (Figure 3B) and EEG-confirmed (Figure 3D) seizures in the preceding 24-hour period. Prior studies evaluating seizure self-forecasting have used self-reported seizure diaries without EEG confirmation. Given the documented difficulties in accurate seizure reporting by patients and caregivers, long-term studies with concurrent objective confirmation of seizures are needed. Newly available devices for continuous monitoring of seizures now make it feasible to conduct such studies.

The lack of predictive association of seizure occurrence with missed medication doses in some of our analyses is most likely due to the rarity of missed medications throughout our dataset and the limited statistical power of rare responses in a survey study. This could highlight a potential selection bias in our data, as our inclusion requirement of good adherence with the electronic survey may preferentially select subjects with personality traits or support systems that also result in good adherence to a medication regimen.

As noted above, we observed strong day-to-day groupings of reported seizures (Figure 4C, SOG, n = 69). We further looked at the data under the condition that a recent seizure had been reported before the survey (Supplemental Figure 1). Under this condition, perceived risk was still the only factor significantly associated with future reported seizure. Importantly, PPV measures at all three scales of perceived risk were significantly higher (Figure 4D).

The OR has been widely accepted as an indicator of test performance in diagnostic studies in medicine[26]. However, OR, which reflects the relative odds change, usually does not provide a direct measure of sensitivity and specificity, which are typically reported in device studies28 and are necessary to establish a method’s usefulness[28]. It should be noted that our derivation of the equations is under the assumption that OR is constant for all possible cutoff values. Otherwise, a closed form mathematical relation between the OR and AUC generally does not exist[26].

The small size of the cohort with concurrent EEG is a limitation in this study. This was caused primarily by the challenges in long-term ambulatory EEG monitoring[41] and the burden on patients of managing and charging multiple devices. Adherence to the e-surveys was not monitored as closely as adherence to wearable and implanted EEG devices during the study, and many patients felt overwhelmed by these requirements and did not complete the e-surveys. Patient burden is a significant challenge in longitudinal studies whether using devices or surveys and incorporating these considerations into the design of new devices and monitoring systems is important.

## 5 Conclusion

This multi-center, prospective study of e-survey results evaluating premonitory factors with future self-reported and EEG-validated seizures has shown that while low mood and high stress are individually associated with future self-reported seizures, they are also significantly associated with previous self-reported seizures. When these recent seizures are included in the statistical model, only perceived seizure risk remains significantly predictive of future self-reported seizures, suggesting these other factors do not contribute independent predictive information. In the small subgroup with concurrent EEG, we did not find evidence to support the ability of patients to self-forecast EEG-confirmed seizures. Reliable self-forecast of seizures would be tremendously helpful to patients, making “pre-emptive treatment”[12] possible without the use of devices, but long-term study in a large cohort with concurrent EEG is needed to assess this possibility.

## Data Availability

Limited data are available upon reasonable request to the authors.

## Appendix

### A. Derivation of converter from OR to AUC

Equation (1) shows a curve on FPR-TPR plot for a given OR. To find the closed form, Equation (2), for the AUC as a function of OR, we may resort to the basic integration equation

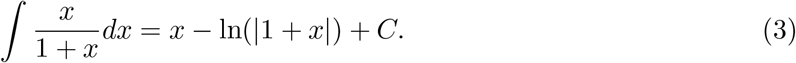

Let FPR be *x* and OR be *γ*, from Equation (1) we have

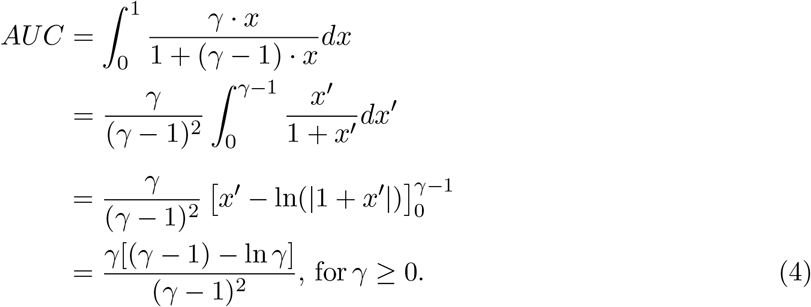

Due to Equation (1), *AUC* = 1*/*2, if *γ* = 1.

## Author contributions

J.C.: Conceptualization, data analysis and composition of the manuscript. I.B.: Conceptualization, data collection, data analysis and manuscript revision. E.N.: Data collection and manuscript revision. P.V.: Data collection, seizure identification and manuscript revision. N.G.: Data collection. P.K.: Data collection. G.A.W: Funding acquisition, data collection and seizure identification. M.P.R: Conceptualization, data collection and manuscript revision. B.H.B: Funding acquisition, conceptualization, data collection and composition of the manuscript

## Acknowledgments

We thank Dr. Mona Nasseri for valuable comments throughout the preparation of the manuscript and Filip Mivalt for providing with MATLAB code of IEA detection for iEEG signals.

## Data availability

Limited data are available upon reasonable request to the authors.

## Supplemental Material

### A. Interictal epileptiform activity (IEA)

In order to assess whether survey responses had any relationship to interictal EEG activity, we also acquired IEA counts and estimated the hourly interictal spike rate from the concurrent EEG. For the RNS device, hourly detection of spike like IEA were stored on the device and then uploaded to a cloud system, where the continuous timestamps of spike hourly counts were then downloaded.

For the subcutaneous EEG signals, we deployed an IEA detector adapted from a spike detector developed for iEEG signals[42]. Signals were preprocessed by computing the root-mean-squared (RMS) envelope and thresholding the signal to exclude high amplitude chewing artifacts. The preprocessed signals were then analyzed using Janca’s algorithm[42]to obtain candidate spikes, which were further selected by imposing upper and lower thresholds on the amplitudes of the candidate spikes to exclude low amplitude background and high amplitude residual myogenic activity. We then computed the hourly spike rates in each channel and took the mean of the hourly spike rates from the two sqEEG channels.

## Supplemental figures

**Supplemental Figure 1:**
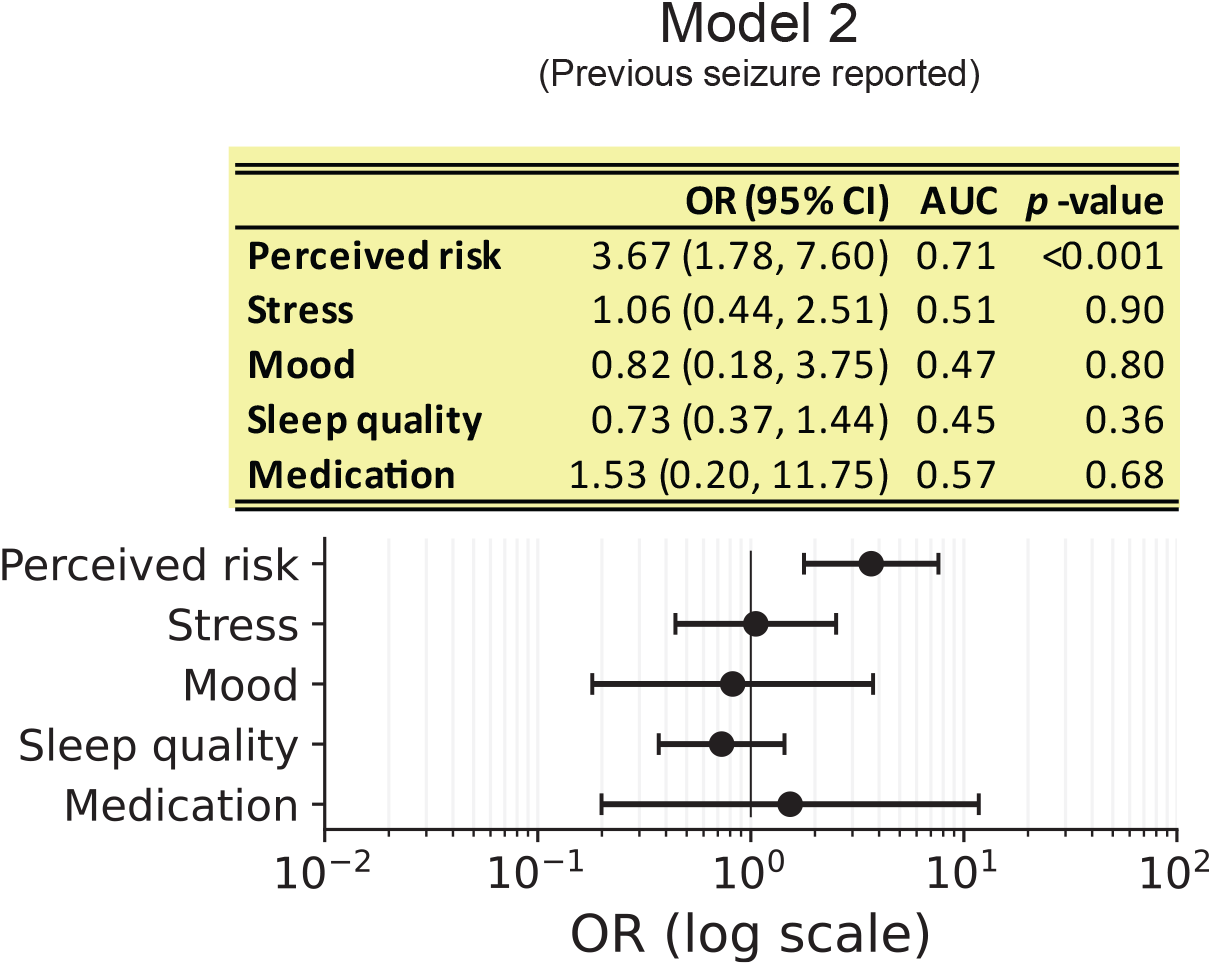
Multivariate regression modeling analysis of reported seizure in next 24 hours in survey only group (SOG, n = 69). The table presents the estimated values of OR, 95% CI, AUC and p-value (Jackknife standard error), corresponding to the error bars shown below. ORs corresponding to the combination of a set of predictors of those in Model 2 (Figure 4B) given that previous seizures (in prior 24 hours) were reported are shown.

**Supplemental Figure 2:**
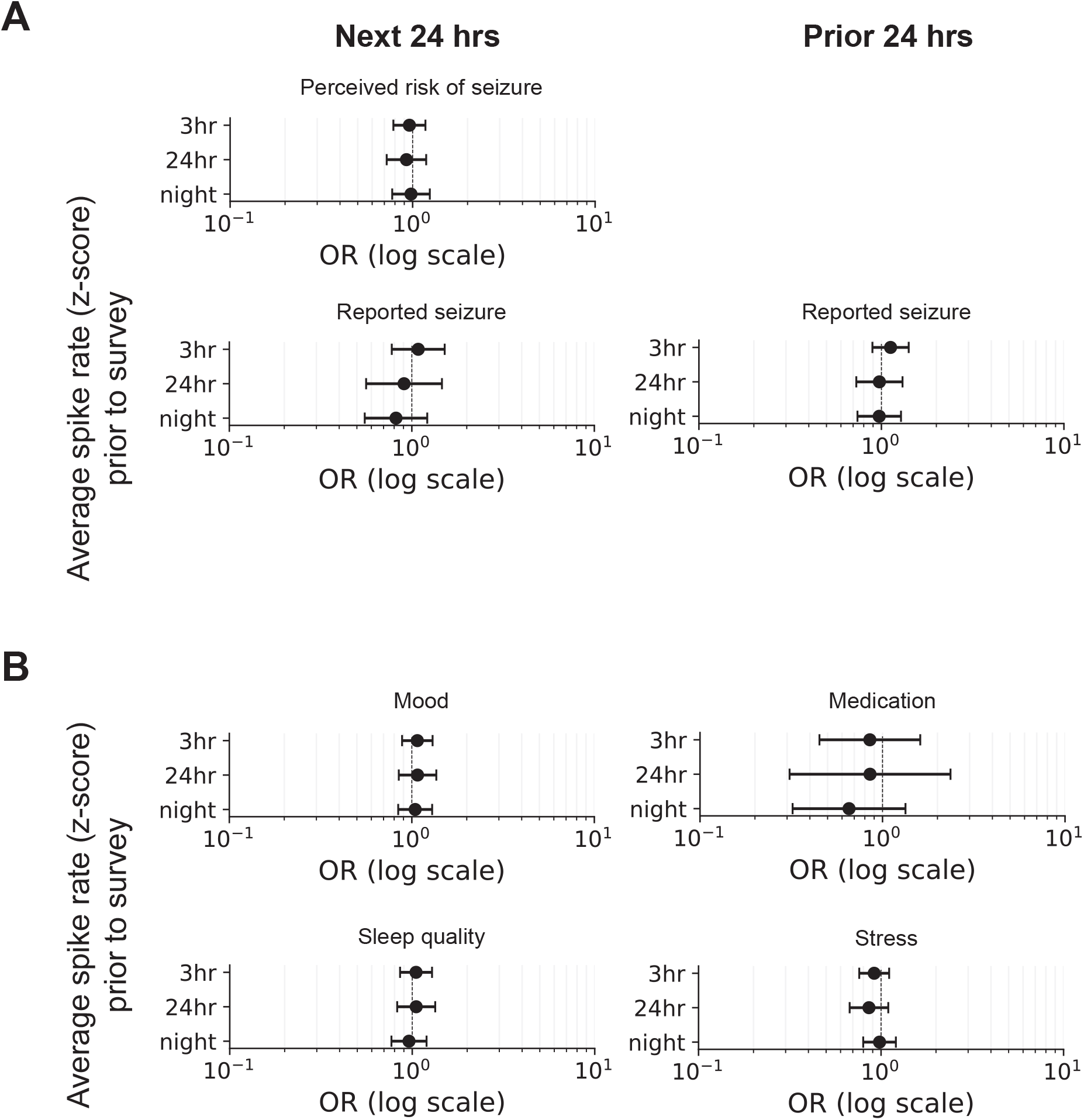
ORs between average spike rates (3 hours, 24 hours or overnight) prior to surveys and factors reported by surveys in CEG (n = 4). (A) ORs between the average spike rates and seizure events. Seizure events include the perceived risk of seizure in next 24 hours and the self-reported seizure either in next 24 hours (left column) or in 24 hours prior to the survey (right column). (B) ORs between the average spike rates and premonitory factors. Error bar: 95% CI. Abbreviation: OR, odds ratio; hrs, hours; CI, confidence interval.

**Supplemental Figure 3:**
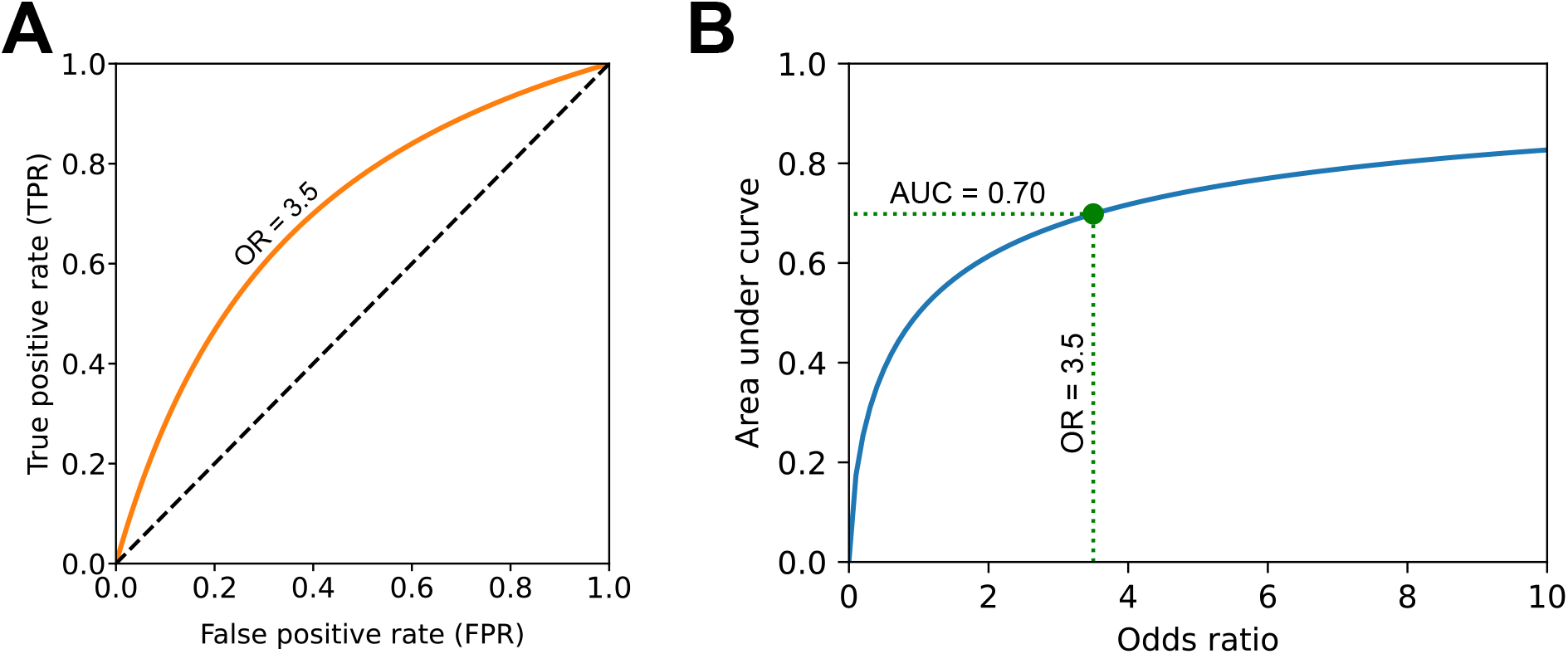
Equal OR curve and the relationship between OR and AUC. (A) An example of equal OR curve from Equation (1), assuming constant OR (e.g., 3.5) for all possible cutoff values. The curve is symmetric in relation to the diagonal *x* + *y* = 1. (B) AUC of equal OR curve as a function of OR from Equation (2). Green dot and the dotted lines indicate the corresponding value of the example shown in Panel A. Note: AUC will be less than 1*/*2 for OR ≤ 1. Abbreviation: OR, odds ratio; AUC, area under curve; TPR, true positive rate; FPR, false positive rate.

## Supplemental tables

**Supplemental Table 1:** Electronic survey questions and conversion of site-specific questions. All participants completed the same survey across three sites, except one participant at KCL. The response of this individual KCL participant is converted. Note: †KCL subject 1 not included in medication analysis. *The survey question asking if a seizure occurred in the past 24 hours was in addition to the separate, self-reported seizure diary. Abbreviation: Q, question; R, response.

|  | Mayo, Seer, KCL (n = 68) | †KCL Subject 1 (n = 1) | Conversion |
| --- | --- | --- | --- |
| **Mood** | Q. "Rate your current mood"<br>R. 5 scales: *Very sad, sad, neutral, happy, very happy* |  | One to one |
| **Stress** | Q. "Are you more or less stressed than you usually are?"<br>R. 3 scales: *Less, as usual, more* | Q. "How much stress do you feel right now"<br>R. 5 scales: *Very relaxed, relaxed, neutral, stressed, very stressed* | • KCL < 25th percentile → Mayo "less"<br>• KCL 25th - 75th percentile → Mayo "as usual"<br>• KCL > 75th percentile → Mayo "more" |
| **Sleep duration** |  | Q. "How many hours did you sleep last night?"<br>R. *Score between 0 and 1* | Discarded question |
| **Sleep quality** | Q. "How did you sleep last night compared to usual?"<br>R. 3 scales: *Worse, as usual, better* | Q. "Rate your sleep quality last night"<br>R. 3 scales: *lower, normal, higher* | One to one |
| **Medication** | Q. "Did you take all of your prescribed epilepsy medication in the last 24 hours?"<br>R. 3 scales: *yes/no/Not Available* |  | KCL subject 1 not included in medication analysis |
| **Perceived risk** | Q. "What do you feel your risk of a seizure is in the next 24 hours?"<br>R. 3 scales: *low, medium, high* |  | One to one |
| **Seizure occurrence** | Q. "Have you had a seizure in the last 24 hours?" *<br>R. 2 scales: *yes/no* |  |  |

**Supplemental Table 2:** Electronic surveys collected by site, type, and analysis group. This table shows cohort and sample sizes for the two cohorts with and without concurrent electrographic seizure logs. Two of the Mayo subjects with concurrent NeuroPace RNS recordings were excluded from CEG (see text).

| Cohort | Survey only group<br>(SOG, n = 69) |  | Concurrent EEG<br>group (CEG, n = 4) |  |
| --- | --- | --- | --- | --- |
|  | Participants | Survey entries | Participants | Survey entries |
| Mayo | 4 | 615 | 2 | 387 |
| KCL | 2 | 262 | 2 | 209 |
| Seer | 63 | 11,713 |  |  |
| Total | 69 | 12,590 | 4 | 596 |

## Footnotes

1 Supplemental Table 3 is not presented in MedRxiv preprint.

